# Epicardial and hepatic fat in Fontan patients is associated with ventricular changes on cardiac MRI

**DOI:** 10.1101/2025.07.08.25331155

**Authors:** Meghan Drastal Ryan, Nicole St. Clair, Christine K. Lee, Rahul H. Rathod

## Abstract

**Background:** Epicardial and hepatic fat are suspected to play a role in cardiac remodeling. The primary objective of this study was to identify the cardiac MRI (CMR) parameters that are associated with indexed epicardial fat volume (EFV*_i_*) and hepatic steatosis in patients after the Fontan operation.

**Methods:** This was a single-center, retrospective analysis of Fontan patients. Epicardial and subcutaneous fat were analyzed with CMR post-processing software, cvi42 (Circle Cardiovascular Imaging, Calgary, Alberta, Canada). Hepatic steatosis was measured with controlled attenuation parameter (CAP) scores via vibration controlled transient elastography.

**Results:** The cohort included 81 patients (64% male, median age 16 years). On univariate analysis, EFV*_i_* correlated with BMI (ρ=0.52, p<0.001), age (ρ=0.48, p<0.001), EDV*_i_* (ρ=0.40, p<0.001), ventricular mass*_i_* (ρ=0.39, p<0.001), subcutaneous fat thickness (ρ=0.31, p<0.01), and inferior vena cava (IVC) flow (ρ=0.31, p<0.05). On multivariable regression analysis, BMI (β 2.16, p<0.001) and EDV*_i_* (β 0.30, p<0.001) were independently associated with EFV*_i_* (R^2^ 0.47). Males had higher median CAP scores than females (222 vs 196 dB/m; p<0.05). On univariate analysis, CAP scores correlated with BMI (ρ=0.53, p<0.001), subcutaneous fat thickness (ρ=0.48, p<0.001), age (ρ=0.35, p<0.01), EFV*_i_* (ρ=0.34, p<0.01), and IVC flow (ρ=0.29, p=0.02). On multivariable analysis, BMI (β 5.8, p<0.001) was independently associated with CAP scores (R^2^=0.28). There were no significant relationships of EFV*_i_* or CAP with adverse clinical events.

**Conclusion:** BMI is a strong predictor of hepatic and epicardial fat distribution. Epicardial fat volume is also associated with ventricular dilation and may play a role in adverse cardiac remodeling.

## Introduction

Medical advancements have led to significant improvements in the survival of Fontan patients.^1,2^ However, there are long-term complications due to the chronically elevated systemic venous hypertension and reduced cardiac output inherent in the Fontan circulation. Not only are patients at risk for ventricular dysfunction, but also complications that affect end-organ function.^1,3^ Further, these effects may be compounded by the cardiometabolic consequences of obesity.^4,5^

While the relationship between obesity and cardiovascular disease is well-defined, the influence of visceral and organ-specific adiposity on cardiovascular health is an emerging area of interest. Due to its pro-inflammatory and fibrotic effects, epicardial fat has been implicated in the development of pathogenic conditions, such as coronary artery disease, arrhythmias, and heart failure in the general population.^6^ Separately, hepatic steatosis has been found to be an independent predictor of early cardiac remodeling and associated with diastolic dysfunction in adult populations.^7,8^ Techniques for measuring epicardial fat volume are relatively new and primarily involve the use of MRI or CT.^6^ Non-invasive techniques for assessing hepatic steatosis have also gained interest in recent years.^9,10^ Controlled attenuation parameter (CAP), a vibration-controlled transient elastography-based technique, provides quantitative prediction of the severity of hepatic steatosis.^10^

Fontan patients have significant rates of obesity, with a prevalence comparable to that of the general population.^11^ Further, obesity in patients after the Fontan operation has been associated with increased morbidity and mortality. ^4,12,13^ Risk factors for the development of hepatic fat in Fontan patients appear to be similar to those of the general population, such as obesity and dyslipidemia.^14^ Although the impact of obesity on Fontan circulation has become an emerging area of interest, there is limited research investigating the influence of epicardial and hepatic fat deposition in this population. Recent work suggests that visceral and epicardial adipose tissue may negatively impact hemodynamics in the Fontan circulation.^15,16^ To further investigate this, we aimed to examine the clinical significance of epicardial and hepatic fat in Fontan patients.

Our primary objective was to identify which clinical factors and cardiac MRI (CMR) parameters were associated with epicardial fat volume indexed to BSA (EFV*_i_*) and hepatic steatosis. Additionally, we sought to determine the relationships between epicardial fat, hepatic fat, and subcutaneous fat. Finally, we studied whether EFV*_i_* and hepatic steatosis were linked to adverse clinical outcomes.

## Methods

### Patient Selection

This was a single-center, retrospective analysis of patients who have undergone the Fontan operation. Fontan patients were included if they had vibration-controlled transient elastography (VCTE) and CMR within one year of each other. Exclusion criteria included growth hormone and steroid use due to concern for confounding hepatic fat distribution. Patients were excluded if there were technically inadequate CMR images or CAP measurements needed for analysis. Significant outliers were also excluded. The study was approved by the Institutional Review Board (IRB) at Boston Children’s Hospital.

### Clinical parameters

Patient demographic and clinical data including cardiac diagnosis, surgical history and laboratory values were abstracted from the medical records. Body mass index (BMI) categories were determined based on CDC growth chart percentiles for patients 2 to 19 years of age and standard adult BMI classifications for patients 20 years and older. BMI classification groups included underweight (<5^th^ percentile or <18.5), normal weight (5^th^ to <85^th^ percentile or 18.5 to 24.9), overweight (85^th^ to <95^th^ percentile or 25 to 29.9), and obese (≥95^th^ percentile or ≥30).

Surgical history data included information on all surgeries and catheterization interventions. Major adverse events were also collected, including history of protein-losing enteropathy (PLE), stroke, thrombus, seizures, pleural effusion, and hemoptysis. Arrhythmia history was compiled through a review of Holter monitors, EP studies and clinic notes; arrhythmic events included atrial fibrillation, atrial flutter, supraventricular tachycardia, non-sustained ventricular tachycardia (≥3 beats lasting <30 s), sustained ventricular tachycardia (≥30 s) and history of cardiac arrest and cardiac rhythm management devices (i.e., pacemakers, defibrillators).

### CMR & VCTE Data Acquisition

CMR studies were performed on a 1.5 Tesla scanner (Philips Healthcare, Best, Netherlands). The details of the CMR protocol have been published previously.^17,18^ Concisely, epicardial fat was measured on an electrocardiogram-gated, steady-state free precession (SSFP) cine image in a short-axis plane from the atrioventricular junction through the apex of the heart. Subcutaneous fat was measured on SSFP axial images at the level of the diaphragm and then confirmed with cross-sectional coronal images. Controlled attenuation parameter (CAP) measurements (dB/m) and liver stiffness measurements (kPA) were measured through vibration-controlled transient elastography (VCTE) using FibroScan^®^ (Echosens, Paris, France). Hepatic fatty infiltration was defined by a CAP score greater than or equal to 215.

### CMR Analysis

CMR data were abstracted from the clinical reports. To account for variations in body size, single ventricle end-diastolic volume (EDV), end-systolic volume (ESV), mass, and stroke volume (SV) were indexed to body surface area (BSA).^19-21^ The indexed values are represented by EDV*_i_*, ESV*_i_*, mass*_i_*, and SV*_i_*, respectively.

Epicardial and subcutaneous fat were analyzed with CMR post-processing software, cvi42 (Circle Cardiovascular Imaging, Calgary, Alberta, Canada). Epicardial fat (EFV) was manually traced with contour tools on consecutive end-systolic short-axis images as shown in Figure 1a. Cine MRI sequences or dynamic, real-time imaging sequences were used to assess cardiac motion and help delineate epicardial fat from external layers of fat and/or scar tissue. EFV was calculated using the density conversion between fat (0.9 g/mL) and myocardial tissue (1.05 g/mL) and then indexed to body surface area (EFV*_i_*).

**Figure 1:**
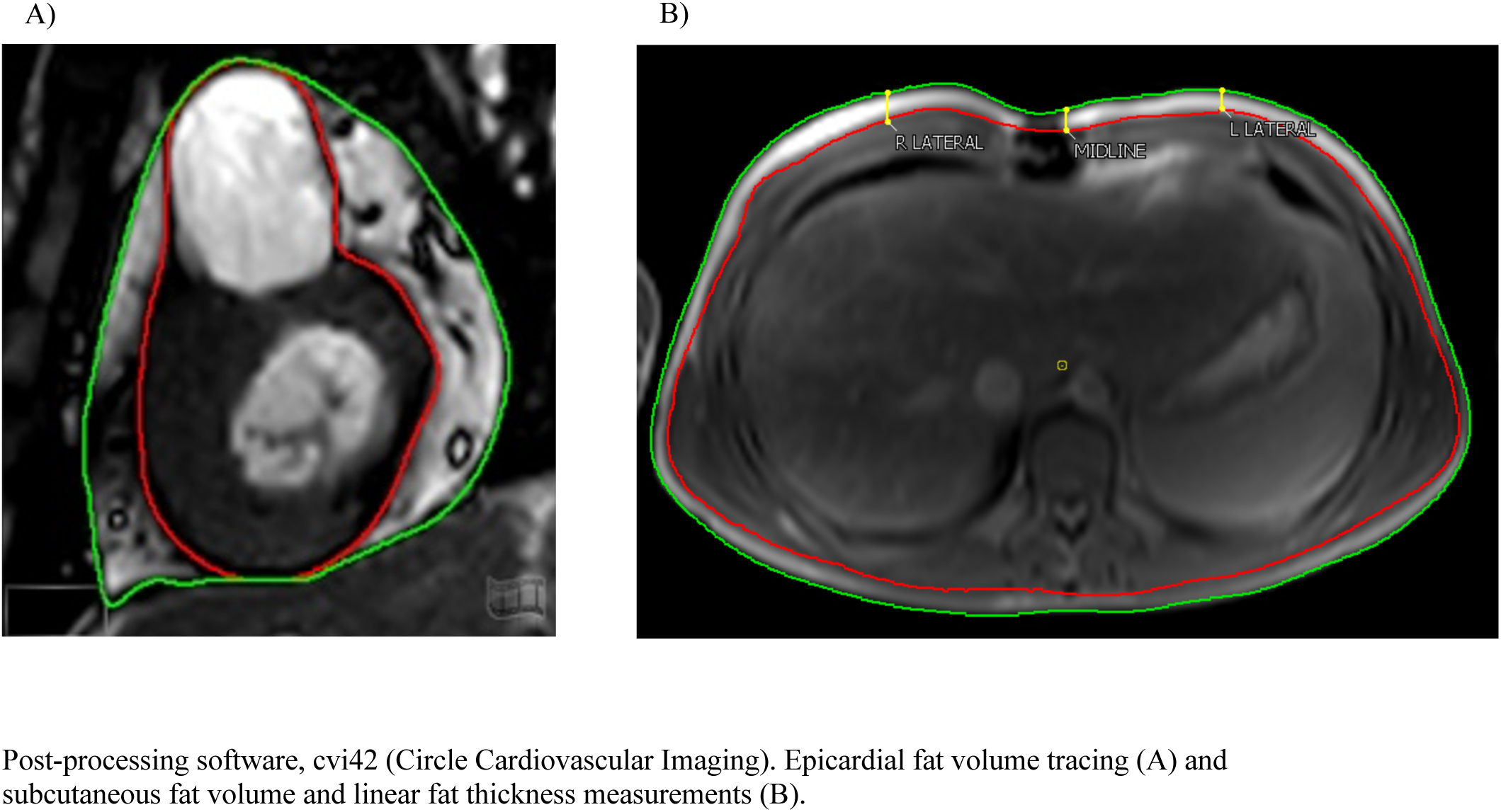
Epicardial and Subcutaneous Fat Analysis.

Subcutaneous fat was manually traced on an axial slice at the level of the diaphragm. Similarly, the density conversion between fat and myocardial tissue was used to define subcutaneous fat volume, which was indexed to BSA (SQFV*_i_*), as shown in Figure 1b. These methods were adapted from Lubert et., 2018 and Lubert et al., 2020.^15,16^ Subcutaneous linear thickness was measured perpendicularly at the midpoint between the outer border of the visceral compartment and midline (spinal reference point), as shown in Figure 1b. The values obtained on either side of the midline were averaged together to find the average subcutaneous linear thickness.

### Statistical Analysis

For data analysis, the Student’s t-test or Wilcoxon rank-sum tests were used to compare continuous variables between two independent groups. Chi-squared test or Fisher’s exact test were used to compare categorical variables. One-way ANOVA tests were used to compare continuous variables between three or more groups. Spearman correlation tests were used to analyze the strength of associations between continuous variables. Multivariable linear regression analyses were used to investigate the associations of EFV*_i_* and CAP scores with imaging and clinical variables. All p-values were two-tailed with a significance level of p < 0.05.

## Results

### Patient Demographics

The study cohort included 81 Fontan patients of which 64% were male (n=52) and 36% were female (n=29). The median age at time of CMR was 16 years [interquartile range: 12 to 19 years]. The median BMI at time of CMR was 21.73 [19.28, 25.04]. The majority of patients were normal weight (n=53, 65%), followed by obese (n=16, 20%), overweight (n=8, 10%), and underweight (n=4, 5%). The patient characteristics of the study population and CMR data stratified by BMI categories are summarized in Table 1.

**Table 1:**
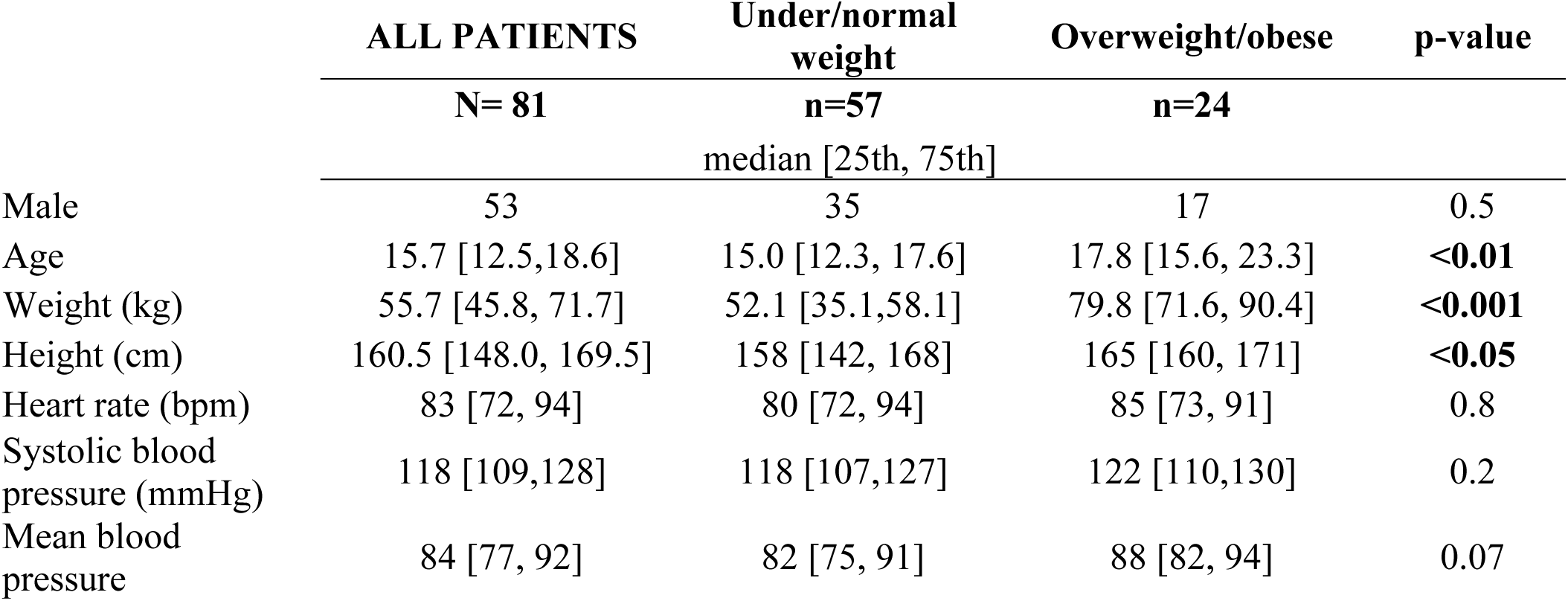
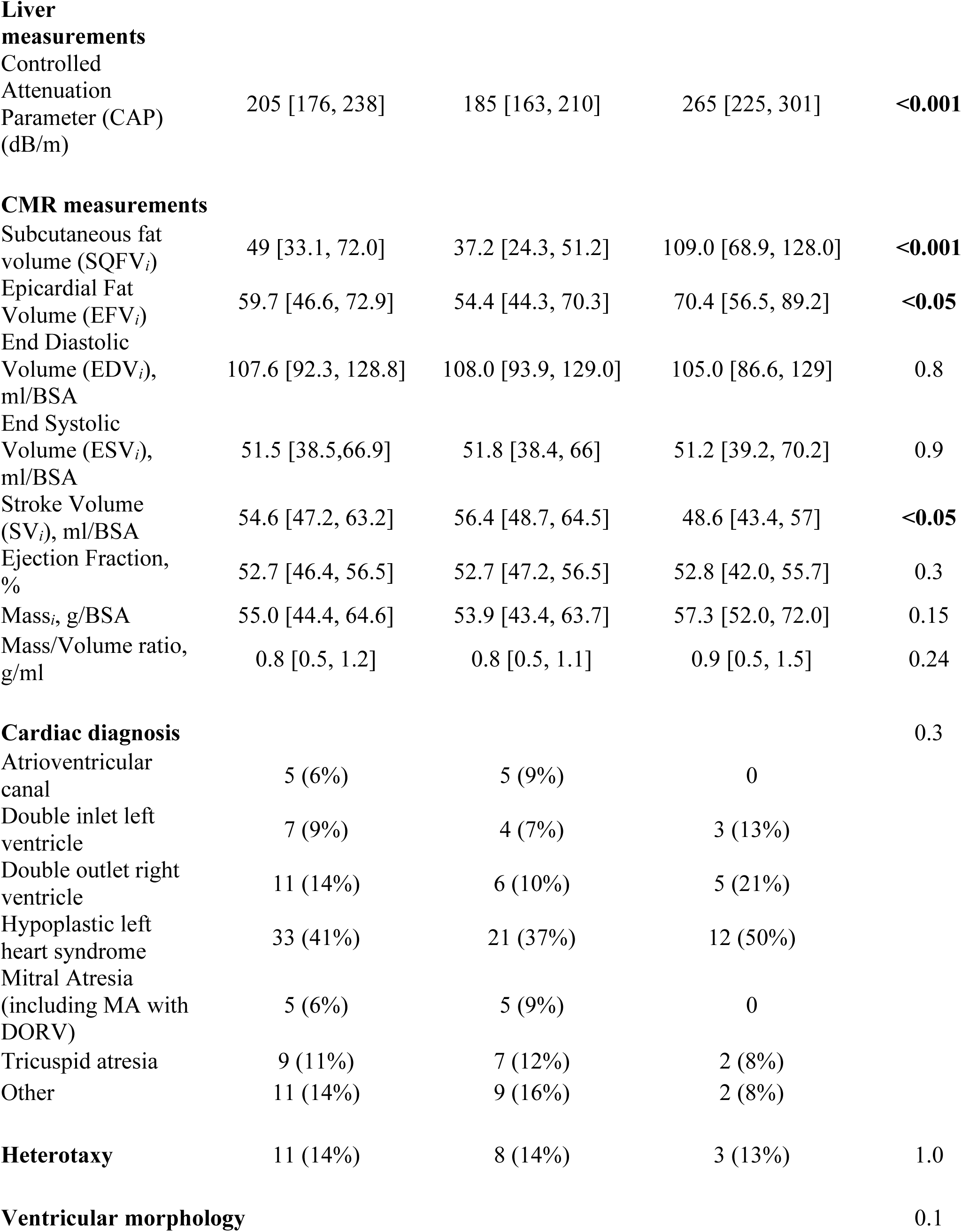
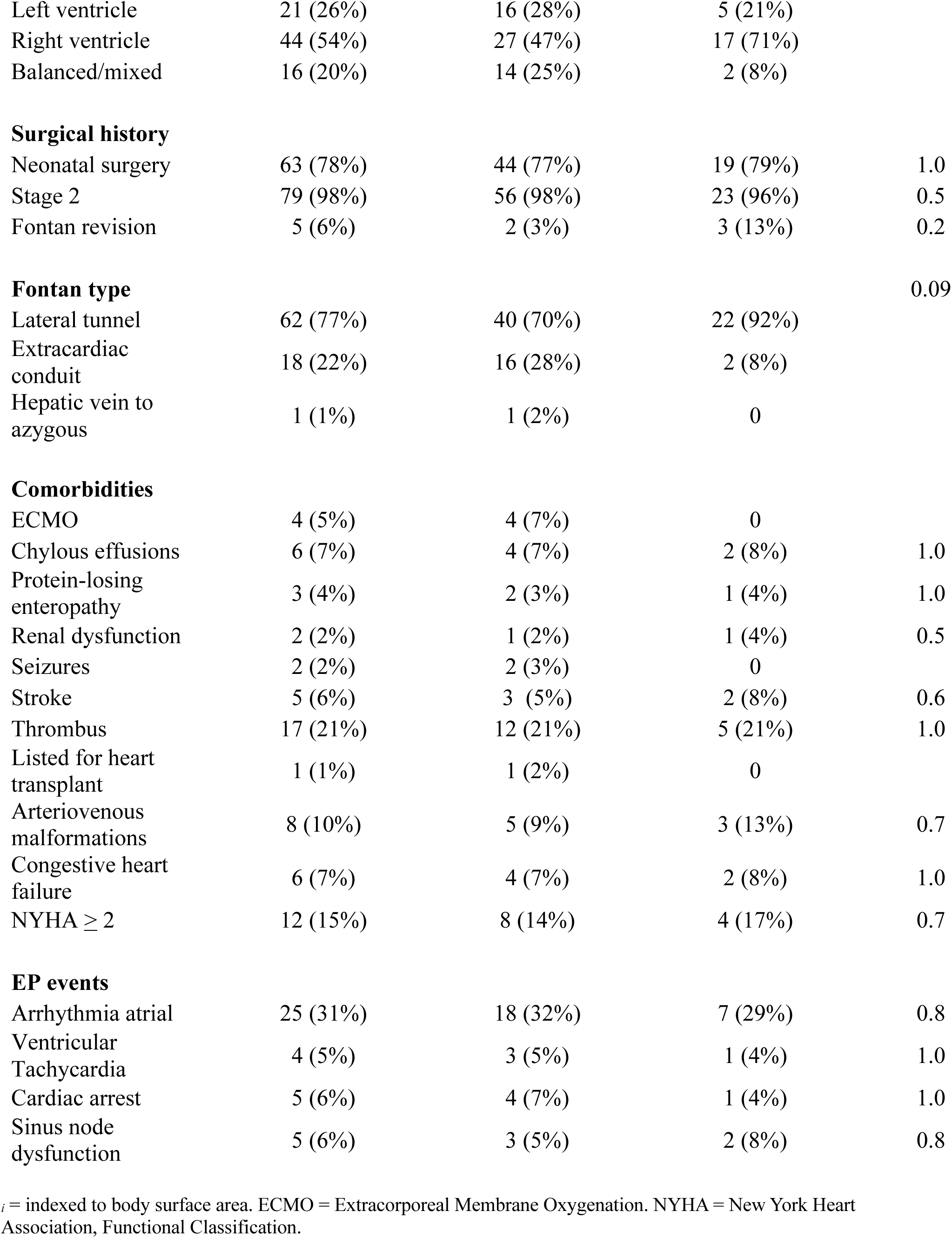
Baseline & Imaging Characteristics.

### Epicardial Fat Volume Analysis

EFV*_i_* demonstrated moderate positive correlations with BMI (ρ=0.52, p<0.001) and age (ρ=0.48, p<0.001). There were weak to moderate correlations between EFV*_i_* and single ventricle (SV) SV ESV*_i_* (ρ=0.42, p<0.001), EDV*_i_* (ρ=0.40, p<0.001), SV mass*_i_* (ρ=0.39, p<0.01), total bilirubin (ρ=0.35, p<0.01), CAP scores (ρ=0.34, p<0.01), subcutaneous fat thickness (ρ=0.31, p<0.01), and inferior vena cava (IVC) flow (ρ= 0.31, p<0.05). EFV*_i_* was weakly correlated with right pulmonary artery flow (ρ=0.24, p<0.05), subcutaneous fat volume (ρ=0.22, p<0.05), and SV stroke volume (SV*_i_*) (ρ=0.22, p<0.05). There was a weak, negative correlation between EFV_i_ and ejection fraction (ρ=-0.34, p<0.01). In multivariable linear regression analysis, BMI (β 2.16, p<0.001) and SV EDV*_i_* (β 0.30, p<0.001) were independently associated with EFV*_i_* (R^2^ 0.47) (see Table 3). There were no statistically significant relationships between EFV*_i_* and major clinical events or adverse outcomes. Univariate and multivariate correlation analyses are summarized in Tables 2 and 3.

**Table 2:** Univariate epicardial fat volume*_i_* correlation analysis.

| Variable | Correlation Coefficient ($\rho$) | p-value (p) |
| --- | --- | --- |
| Body mass index | 0.52 | <0.001 |
| Age | 0.48 | <0.001 |
| End systolic volume<sub>i</sub> | 0.42 | <0.001 |
| End diastolic volume<sub>i</sub> | 0.40 | <0.001 |
| Ventricular mass<sub>i</sub> | 0.39 | <0.01 |
| Total bilirubin | 0.35 | <0.01 |
| CAP scores | 0.34 | <0.01 |
| Subcutaneous fat thickness | 0.31 | <0.01 |
| Inferior vena cava flow | 0.31 | <0.05 |
| Right pulmonary artery flow | 0.24 | <0.05 |
| Subcutaneous fat volume | 0.22 | <0.05 |
| Stroke volume<sub>i</sub> | 0.22 | <0.05 |
| Ejection fraction | -0.34 | <0.01 |

<sub>i</sub> = indexed to body surface area. CAP = Controlled Attenuation Parameter.

**Table 3:** Multivariate indexed epicardial fat volume linear regression analysis.

| Variable | Beta Coefficient ($\beta$) | p-value (p) |
| --- | --- | --- |
| Body mass index | 2.16 | <0.001 |
| End diastolic volume<sub>i</sub> | 0.30 | <0.001 |

<sub>i</sub> = indexed to body surface area.

### Hepatic Steatosis Analysis

The median CAP score was 205 dB/m [176, 238]. Nearly 40% of patients (n= 32) had CAP scores suggesting the presence of hepatic fatty infiltration. Specifically, 24 patients (30%) had CAP scores ranging from 215 to 300 dB/m, representing mild to moderate steatosis, and 8 patients (10%) had CAP scores >300 dB/m suggestive of severe steatosis. Males had significantly higher median CAP scores than females (222 vs 196 dB/m; p<0.05). CAP scores demonstrated strong to moderate, positive correlations with BMI (ρ=0.53, p<0.001), subcutaneous fat thickness (ρ=0.48, p<0.001), subcutaneous fat volume (ρ=0.47, p<0.001), and age (ρ=0.35, p<0.01). There were weak, positive correlations between CAP scores and IVC flow (ρ=0.29, p<0.05). Following multivariable analysis, only BMI (β 5.8, p<0.001) was independently associated with CAP scores (R^2^=0.28). No statistically significant associations between CAP scores and adverse clinical events. Univariate and multivariate correlation analyses are summarized in Tables 4 and 5.

**Table 4:** Univariate controlled attenuation parameter (CAP) score correlation analysis.

| Variable | Correlation Coefficient ($\rho$) | p-value (p) |
| --- | --- | --- |
| Body mass index | 0.53 | <0.001 |
| Subcutaneous fat thickness | 0.48 | <0.001 |
| Subcutaneous fat volume | 0.47 | <0.001 |
| Age | 0.35 | <0.01 |
| Epicardial fat volume<sub>i</sub> | 0.34 | <0.01 |
| Inferior vena cava flow | 0.29 | <0.05 |

<sub>i</sub> = indexed to body surface area.

**Table 5:** Multivariate controlled attenuation parameter (CAP) score linear regression analysis.

| **Variable** | **Beta Coefficient ($\beta$)** | **p-value (p)** |
| --- | --- | --- |
| Body mass index | 5.8 | <0.001 |

## Discussion

This study demonstrates that BMI is a strong predictor of both hepatic and epicardial fat distribution. Epicardial fat volume is independently associated with ventricular dilation. Epicardial and hepatic fat may play an important role in cardiac remodeling and have implications regarding cardiac function.

### BMI-Related Outcomes in Fontan Patients

In our patient population, nearly one third of Fontan patients were overweight or obese, which is consistent with earlier findings on the prevalence of elevated BMIs in other Fontan cohorts.^11^ Previous work has established the association between increased BMI and poor outcomes in Fontan patients. Fontan patients with elevated BMI have worse hemodynamics, such as decreased peak oxygen consumption, increased Fontan pressure, and pulmonary capillary wedge pressure.^13^ Further, Fontan patients with an elevated BMI are at increased risk for heart failure hospitalization and thromboembolic complications.^13^ A recent large, multi-center study (n=3,778, 35 centers) from the Fontan Outcomes Registry using CMR Examinations found that elevated BMI was associated with increased end diastolic volume and ventricular mass. Fontan patients with obesity had both higher ventricular end diastolic and Fontan pressures by cardiac catheterization as well. ^12^

### Role of Visceral Fat Distribution: Epicardial Adipose Tissue

As suggested in the current study, visceral fat distribution may play a more significant role in ventricular adaption than BMI alone. In other patient populations, such as adult patients with heart failure with preserved ejection fraction and pulmonary hypertension, epicardial adipose tissue has been associated with increased risk of morality and worse survival.^22,23^ Our study builds on contemporary work highlighting the growing recognition regarding the clinical significance of epicardial fat in the Fontan population. Lubert et al., 2018 found that Fontan patients have higher epicardial fat volume compared to repaired Tetralogy of Fallot patients. In addition, the researchers observed a weak, inverse correlation between EFV and both lower systemic ventricle ejection fraction and cardiac output in Fontan patients.^16^ A cross-sectional analysis investigating visceral adiposity and Fontan hemodynamics as measured by cardiac catheterization demonstrated that epicardial fat volume was independently associated with pulmonary vascular resistance.^15^ The present work builds on this growing body of literature, signifying that epicardial fat volume may play a role in deleterious cardiac modeling as evidenced by changes to end-diastolic volume. Previous research has suggested that changes to ventricular volumes have clinically significant, negative implications on the Fontan circulation. Specifically, a large cohort of Fontan patients demonstrated that increased ventricular dilation as measured by EDV on cardiac MRI was the strongest independent predictor of death or transplant listing in Fontan patients.^24^ Further, our study further validates novel methods for measuring epicardial fat volume using CMR post-processing software.^15,16^

Several mechanisms have been proposed to explain how epicardial adipose tissue may drive alterations in cardiac structure and function. Specifically, epicardial fat contributes to the development of cardiovascular disease through altered gene expression leading to a pro-inflammatory and profibrotic profile as well as interferences in glucose and lipid metabolism.^6^ The metabolic activity of local adipose tissue may drive an inflammatory response leading to the release of cytokines that contribute to endothelial dysfunction, myocardial fibrosis, and ventricular remodeling.^25^

### Role of Visceral Fat Distribution: Hepatic Steatosis

As alluded to earlier, this study not only focuses on epicardial adipose tissue as a measure of visceral fat, but also hepatic steatosis. Katz et al., 2021 examined the prevalence and associations of hepatic steatosis as measured by abdominal MRI in Fontan patients and found that hepatic steatosis was present in approximately 10% of the study population. Hepatic steatosis was associated with increased BMI and lower HDL cholesterol.^14^ As previously demonstrated, obesity remained a strong, independent predictor of the presence of hepatic steatosis in Fontan patients. However, in contrast to the aforementioned study, hepatic steatosis, as defined by a CAP score of 215-300 dB/m, was present in greater than 40% of patients in our cohort. To our knowledge, this is the first analysis of hepatic fat using CAP scores in Fontan patients.

While hepatic steatosis has previously been linked to early changes in cardiac structure and function, such LV remodeling, diastolic dysfunction, and reduced systolic function, the presence of obesity appears to attenuate this relationship.^26^ In the current study, only BMI remained significant in the final model.

### Limitations

Inherent limitations are introduced by design, as a single-center, retrospective, cross-sectional study. While referral bias may have led to an overrepresentation of more symptomatic patients undergoing CMR, this is mitigated by our center’s standardized practice of performing CMRs in all Fontan patients once old enough to tolerate imaging protocols. Similarly, in recent years, our center has also started to routinely perform VCTE in patients seen in the Fontan at Boston multidisciplinary clinic. We excluded patients who did not have a CMR and VCTE within 1 year of each other, which may have limited the sample size. Further, the limited number of patients who experienced adverse events may have contributed to the absence of significant associations observed, highlighting the need to for validation in larger studies.

### Clinical Significance & Future Directions

This study further contributes to the growing body of evidence highlighting the detrimental effects of obesity on the Fontan circulation. Nutritional and exercise-related changes aimed at normalizing BMI may modulate these effects. In our analysis, BMI was the strongest independent predictor of both epicardial and hepatic fat. By focusing on interventions to address elevated BMIs, visceral fat may also be improved.

Independent of their effects on weight loss and blood glucose control, sodium-glucose cotransporter-2 (SGLT2) inhibitors and receptor agonists have been associated with reduced morbidity and mortality in adult patients with heart failure.^27^ Consequently, these medications have garnered interest in the Fontan patient population.^28^ Further, glucagon-like peptide-1 (GLP-1) have demonstrated improved cardiovascular outcomes in adult patients with heart failure and obesity.^29^ Interestingly, both SGLT2 inhibitors and glucagon-like peptide-1 (GLP-1) receptor agonists have also been shown to cause significant reductions in epicardial adipose tissue in patients with Type 2 Diabetes that may be independent of weight loss.^30,31^

Furthermore, it has been postulated that the pro-fibrotic and inflammatory effects of visceral adipose tissue, such as epicardial fat, may be modifiable and potentially reversible.^32^ Epicardial adipose tissue may serve as a promising target for future therapeutic interventions. However, additional research is needed to further establish the clinical significance of visceral fat measures, particularly in the Fontan patient population.

## Conclusion

Increased BMI is strongly associated with visceral fat distribution. Epicardial fat was found to be an independent predictor of ventricular dilation and may be a contributing factor in detrimental cardiac remodeling. Further research is needed to understand the clinical implications of visceral adipose in Fontan patients and potential therapeutic targets.

## Data Availability

Data is available upon request.

## Acknowledgements

This work was supported by grants from the Evan’s Heart Fund and the Lovejoy Research and Education Fund.

